## Supplementary material for "Adolescent choices and caregiver roles: Understanding individual and interpersonal influences on sexual decision-making in South Africa": S1_FGD Guide Adolescents

### FOCUS GROUP GUIDE: ADOLESCENTS (ENROLMENT VISIT)

An observational prospective study evaluating the feasibility of enrolling adolescents and assessing the uptake of essential health services within an adolescent friendly clinical trial setting

***Introduction and Ground Rules***

1. Obtain written informed consent first from all participants, before any data are collected.

2. Interviewer to introduce self. Thank you for taking the time to meet with us today. Our names are [*insert names*] __________ and we would like to talk to you about participation of adolescents in HIV vaccine trials. We are doing this project to understand the behavior of adolescents and how we might help you with sexual decision making. We want you to be as open and honest when answering. There are no right or wrong answers in this discussion. Please feel free to tell us what you think, regardless of whether you agree or disagree with what you hear. We don’t expect everyone to share the same opinions. It is very important that we hear all your opinions.

3. Interviewer to explain the ground rules and terms of confidentiality for the focus group discussion:

    a. The participant does not have to answer any question they do not want to.
    b. The information you share will be handled in confidence. (in secret)
    c. When we report back on the information collected in this discussion, your comments will not be able to be linked to you specifically.
    d. We ask that you also agree not to share anything discussed in this room with others.

4. The discussion should take between one and two hours. We will have a break somewhere in the middle.

5. Interviewer to inform the participants that the focus group discussion will be tape recorded to make sure that all themes are captured. Turn the tape on and ask for verbal permission again to tape record, while the tape is running to verbally capture consent (this is a double check against the written consent). We will be recording the session because we don’t want to miss any of your comments. Although one of us may take some notes while we talk, we can’t write fast enough to get everything down on paper. As we are recording, please try to speak loudly so that we don’t miss your comment

***Questions***

- *In your opinion, what do you think influences your decisions on sexual and reproductive health?*
- *In your opinion, who influences you regarding the decisions that you make on sexual and reproductive health?*
- *Can you tell us about where you learn about sexual and reproductive health?*
- *Can you describe to us why you would agree to take part in a vaccine trial?*
- *Can you describe to us why you would not agree to take part in a vaccine trial?*
- *Can you describe to us at what age you should make your own decisions about your sexual activity?*
- *How do you think we should inform adolescents about risky sexual behaviour?*
- *Can you tell us what measures we could put in place to encourage adolescents to access clinic services for contraception?*
- *Can you describe to us what could encourage you to attend all study visits?*
- *Can you describe to us why you may miss or avoid clinic visits?*
- *Can you describe any recommendations on how we could encourage adolescents to speak honestly about their sexual behaviours?*

***Any other comments***

Are there any final thoughts you have about adolescents, their sexual behavior or participation in vaccine trials?

***End of session***

Now we have come to the end of our discussion. Thank you for your active participation. If you have any questions about your study participation, please contact us. Thank you.

***Time ended (HHMM):***
