## Supplementary material for "Adolescent choices and caregiver roles: Understanding individual and interpersonal influences on sexual decision-making in South Africa": S3_COREQ checklist

**Consolidated criteria for reporting qualitative studies (COREQ): 32-item checklist for manuscript in title:**

| No | Item | Guide questions/description | Response |
|---|---|---|---|
| **Domain 1: Research team and reflexivity** | | | |
| Personal Characteristics | | | |
| 1. | Interviewer/facilitator | Which author/s conducted the interview or focus group? | Done<br>It is mentioned in the method lines 169 to 176 |
| 2. | Credentials | What were the researcher's credentials? *E.g. PhD, MD* | Done<br>It is mentioned in the method lines 169 to 176 |
| 3. | Occupation | What was their occupation at the time of the study? | Done<br>It is mentioned in the method lines 169 to 176 |
| 4. | Gender | Was the researcher male or female? | Done<br>It is mentioned in the method lines 169 to 176 |
| 5. | Experience and training | What experience or training did the researcher have? | Done<br>It is mentioned in the method line 171 |
| Relationship with participants | | | |
| 6. | Relationship established | Was a relationship established prior to study commencement? | No |
| 7. | Participant knowledge of the interviewer | What did the participants know about the researcher? e.g. *personal goals, reasons for doing the research* | Participants and interviewers maintained professional relationships up until the end of the study, mentioned in methods, lines174-176 |
| 8. | Interviewer characteristics | What characteristics were reported about the interviewer/facilitator? e.g. *Bias, assumptions, reasons and interests in the research topic* | None |
| **Domain 2: study design** | | | |
| Theoretical framework | | | |
| 9. | Methodological orientation and Theory | What methodological orientation was stated to underpin the study? *e.g. grounded theory, discourse analysis, ethnography, phenomenology, content analysis* | *thematic analysis*<br>It is mentioned in the method section, lines 189 to 194 |
| Participant selection | | | |
| 10. | Sampling | How were participants selected? *e.g. purposive, convenience, consecutive, snowball* | *Convenience*<br>It is mentioned in the method section, lines 151 to 153 |
| 11. | Method of approach | How were participants approached? e.g. *face-to-face, telephone, mail, email* | *In-person*<br>It is mentioned in the method section, lines 156 |
| 12. | Sample size | How many participants were in the study? | 17 adolescents and 19 caregivers<br>It is mentioned in the results section, lines 215 & 223 |
| 13. | Non-participation | How many people refused to participate or dropped out? Reasons? | 0 |

| | | | |
|---|---|---|---|
| | | | Not mentioned, assumed by sampling technique used. |
| Setting | | | |
| 14. | Setting of data collection | Where was the data collected? e.g. *home, clinic, workplace* | *Research centre* It is mentioned in the method section, line 137 |
| 15. | Presence of non-participants | Was anyone else present besides the participants and researchers? | 0 Not mentioned, assumed by sampling technique used. |
| 16. | Description of sample | What are the important characteristics of the sample? *e.g. demographic data, date* | Done It is mentioned in the method lines 136 to 153 |
| Data collection | | | |
| 17. | Interview guide | Were questions, prompts, guides provided by the authors? Was it pilot tested? | Done It is mentioned in the method lines 163 to 169 |
| 18. | Repeat interviews | Were repeat interviews carried out? If yes, how many? | No Not mentioned |
| 19. | Audio/visual recording | Did the research use audio or visual recording to collect the data? | Yes It is mentioned in the method section, lines 184 to 187. |
| 20. | Field notes | Were field notes made during and/or after the interview or focus group? | Yes It is mentioned in the method section, lines 184 to 187. |
| 21. | Duration | What was the duration of the interviews or focus group? | It is mentioned in the method section, lines 160 to 161. |
| 22. | Data saturation | Was data saturation discussed? | No Limitations mentioned, lines 425 to 430. |
| 23. | Transcripts returned | Were transcripts returned to participants for comment and/or correction? | No Not mentioned |
| **Domain 3: analysis and findings** | | | |
| Data analysis | | | |
| 24. | Number of data coders | How many data coders coded the data? | There was one coder and two reliability checkers. Mentioned in methods section, lines 189 to 192. |
| 25. | Description of the coding tree | Did authors provide a description of the coding tree? | Mentioned lines 189 to 192, which informed theme levels provided in Table 1. |
| 26. | Derivation of themes | Were themes identified in advance or derived from the data? | Mentioned lines 189 to 192, which informed theme levels provided in Table 1. |
| 27. | Software | What software, if applicable, was used to manage the data? | It is mentioned in the method section line 189 |
| 28. | Participant checking | Did participants provide feedback on the findings? | No Not mentioned |
| **Reporting** | | | |
| 29. | Quotations presented | Were participant quotations presented to illustrate the themes / findings? Was each quotation identified? e.g. *participant number* | Done |
| 30. | Data and findings consistent | Was there consistency between the data presented and the findings? | Done |
| 31. | Clarity of major themes | Were major themes clearly presented in the findings? | Done |

| | | | |
|---|---|---|---|
| 32. | Clarity of minor themes | Is there a description of diverse cases or discussion of minor themes? | Table 1 shows the socioecological model (SEM) level, and the themes that emerged for the two SEM levels |
